# Inflammatory protein profiling shows limited ability to distinguish sepsis from sepsis mimics

**DOI:** 10.64898/2026.10.10.26365202

**Authors:** Daisuke Kasugai, Hironoshin Onizuka, Atsushi Numaguchi, Takanori Yamamoto, Noritoshi Kato, Remi Sumiyoshi, Shoichi Fukui, Tomohiro Koga, Atsushi Kawakami, Wataru Fujii, Yutaka Kawahito, Shoichi Maruyama, Teppei Shimamura

**Author notes:** **Corresponding author** Daisuke Kasugai, MD, PhD, Department of Emergency and Critical Care Medicine, Nagoya University Graduate School of Medicine, 65 Tsurumai-cho, Showa-ku, Nagoya 466-8550, Japan.

## Abstract

Sepsis mimics resemble sepsis clinically but may require different treatments. We evaluated whether inflammatory protein profiles could distinguish these conditions in a retrospective case-control study using 4 prospective Japanese cohorts. The study included 34 adults with sepsis and 46 with sepsis mimics. We measured 92 proteins using Olink, assessed between-group differences with false discovery rate (FDR) adjustment, and evaluated 3 prediction models using repeated nested cross-validation. No protein met the FDR threshold; the lowest FDR-adjusted P value was 0.345. The primary elastic-net model had an area under the receiver operating characteristic curve of 0.442 (95% confidence interval, 0.315–0.574), based on cross-validation predictions averaged for each participant. Random forest and gradient boosting also showed no clear evidence of discrimination above chance. Confidence intervals included chance performance in both the all-serum and quality-control–Pass sensitivity analyses. In this cohort, a single measurement of an inflammation-focused protein panel showed limited discrimination between sepsis and sepsis mimics. Integration with clinical information and longitudinal measurements warrants further investigation.

## Introduction

Sepsis is defined as life-threatening organ dysfunction caused by a dysregulated host response to infection.[1] The Global Burden of Disease Study estimated that 48.9 million cases of sepsis and 11.0 million sepsis-related deaths occurred worldwide in 2017.[2] Early recognition and timely treatment of sepsis are therefore major priorities in emergency and critical care. However, some patients initially present with fever, shock, coagulopathy, and multiorgan dysfunction typical of sepsis, although a nonseptic condition is ultimately identified as the principal cause.[3–6] In a retrospective study of 4 Swedish intensive care units, approximately 25% of 2,664 patients admitted with presumed sepsis were classified as having sepsis mimics according to predefined infection criteria.[3] Conditions such as atypical hemolytic uremic syndrome, secondary hemophagocytic lymphohistiocytosis, and thrombocytopenia, anasarca, fever, reticulin fibrosis or renal dysfunction, and organomegaly syndrome can produce organ dysfunction resembling sepsis but may require complement inhibition, immunosuppressive treatment, or plasma exchange rather than antimicrobial treatment alone.[7–9] Distinguishing sepsis from sepsis mimics early enough to guide appropriate treatment is therefore clinically important.[5,6]

Conventional inflammatory biomarkers, including C-reactive protein, procalcitonin, and interleukin-6, have long been used in the evaluation of sepsis.[10] However, single biomarkers show only moderate ability to distinguish infectious from noninfectious inflammation in heterogeneous critically ill populations.[3,11,12] Moreover, sepsis encompasses diverse host responses, making it difficult for a single marker to capture the biological differences relevant to diagnosis. Host-response-based diagnostics and multiplex omics analyses have recently been developed to address this limitation. Blood transcriptomic and proteomic profiles have been used to distinguish infection from noninfectious inflammation,[13] and the proximity extension assay enables simultaneous relative quantification of multiple proteins using a small sample volume.[14] Nevertheless, the extent to which inflammatory protein profiles can distinguish sepsis from a broad range of severe sepsis mimics remains unclear.

We therefore used proximity extension assay-based targeted proteomics to explore the ability of inflammatory protein profiles to distinguish sepsis from sepsis mimics. The primary analysis compared sepsis with the pooled sepsis-mimic group, and secondary analyses compared sepsis with each sepsismimic subgroup. We evaluated both individual proteins and multivariable discrimination models.

## Methods

### Study design and reporting

This retrospective case-control study used stored biospecimens and clinical data from 4 prospective cohorts in Japan. As an exploratory secondary analysis, we evaluated the ability of circulating inflammatory protein profiles to distinguish sepsis from sepsis mimics. The sepsis-mimic group included coronavirus disease 2019, noninfectious acute respiratory distress syndrome, atypical hemolytic uremic syndrome, secondary hemophagocytic lymphohistiocytosis, and thrombocytopenia, anasarca, fever, reticulin fibrosis or renal dysfunction, and organomegaly syndrome. The report was prepared with reference to the Standards for Reporting Diagnostic Accuracy Studies 2015 statement and the Strengthening the Reporting of Observational Studies in Epidemiology statement.[15,16] Reporting of the multivariable prediction analyses was also informed by TRIPOD+AI.[17]

### Source cohorts and participants

Adults aged 18 years or older were eligible if stored serum or plasma and the clinical information required for the analysis were available. Participants were selected according to the availability of stored biospecimens, resulting in approximately 10 participants per sepsis-mimic subgroup. A larger sepsis group was included to improve the balance of the primary comparison and represent a range of organ dysfunction severity.

Participants with sepsis, coronavirus disease 2019 (COVID-19), or noninfectious acute respiratory distress syndrome (ARDS) were selected from the cohort of the AESCULAPIUS multicenter prospective observational study, coordinated by Nagoya University (approval number 2022-0211). Sepsis was defined as suspected infection accompanied by an acute increase in the Sequential Organ Failure Assessment score of at least 2 points.[1] COVID-19 was diagnosed based on SARS-CoV-2 testing and compatible clinical and imaging findings. Although COVID-19 can fulfill Sepsis-3 criteria, it was classified as a sepsis mimic in this study because it is a pathogen-defined syndrome requiring disease-specific management, and prior studies have reported distinct clinical phenotypes and host-response/proteomic features compared with bacterial sepsis.[18–20] Noninfectious ARDS was defined according to the Berlin Definition, with neither an identified causative pathogen nor an evident extrapulmonary focus of infection.[21] Participants with atypical hemolytic uremic syndrome (aHUS) were drawn from the prospective “National research of atypical hemolytic uremic syndrome (aHUS)” cohort (Nagoya University approval number 2019-0433), in which final diagnoses were centrally adjudicated by aHUS experts based on the Japanese atypical hemolytic uremic syndrome clinical guide 2023.[22] Participants in the secondary hemophagocytic lymphohistiocytosis (HLH) group were drawn from a prospective cohort approved by the Ethics Committee of Kyoto Prefectural University of Medicine (approval number ERBC-1618). Patient selection in the parent cohort used the 2016 EULAR/ACR/PRINTO classification criteria for macrophage activation syndrome.[23] Participants with thrombocytopenia, anasarca, fever, reticulin fibrosis or renal dysfunction, and organomegaly (TAFRO) syndrome were drawn from the prospective study “Biomarker analysis in Castleman’s disease, TAFRO syndrome, and related diseases” (University Hospital Medical Information Network registration number UMIN000034188). TAFRO syndrome was diagnosed using the 2015 criteria and severity classification and the 2019 updated criteria. [24,25] Diagnostic assessors determined the diagnoses without knowledge of the Olink results.

### Ethics

All procedures in the present study were performed in accordance with the Declaration of Helsinki and the Ethical Guidelines for Medical and Biological Research Involving Human Subjects issued by the Japanese government. Each parent cohort received approval from the responsible institutional ethics committee, and written informed consent was obtained from all participants or their legal guardians in accordance with its approved protocol. The present secondary analysis used existing specimens and data within the scope of those approvals; an opt-out procedure was additionally used in accordance with these guidelines. The Ethics Review Committee of Nagoya University Graduate School of Medicine approved the secondary analysis (approval number 2023-0314).

### Clinical data

Clinical information included age, sex, disease group, survival status at hospital discharge, Sequential Organ Failure Assessment score, and the inflammatory, coagulation, and immune-related laboratory measurements available in each cohort.

### Biospecimens and targeted proteomic assay

Stored serum samples were used for participants from AESCULAPIUS (n = 51), aHUS (n = 8), and TAFRO (n = 10), whereas stored EDTA plasma samples were used for participants with secondary HLH (n = 11). Targeted proteomic profiling was performed by the Azenta Corporation Gene Analysis Center using the Olink Target 96 Inflammation panel and the proximity extension assay. Samples were analyzed on a single plate according to the manufacturer’s instructions and the testing laboratory’s standard operating procedure. Results were reported as Normalized Protein eXpression (NPX), a relative quantity on a log2 scale. Each sample received 4 internal controls. A deviation of less than 0.3 NPX from the median internal-control value was used as the sample quality criterion. Sample-level quality-control (QC) status was obtained from the laboratory report; samples flagged as Warning were retained in the primary analysis and assessed in a separate sensitivity analysis. The analysis included 92 proteins after excluding internal-control assays. Laboratory assay-performance metrics for the full 88-sample analytical batch are provided in Supplementary Table S4.

### Outcomes

The primary outcome was the ability of the 92-protein inflammatory profile to distinguish sepsis from the pooled sepsis-mimic group. Secondary outcomes were pairwise discrimination between sepsis and each sepsis mimic: COVID-19, noninfectious ARDS, aHUS, secondary HLH, and TAFRO syndrome.

### Sample size and exploratory power assessment

This exploratory secondary analysis used the available specimens, and no formal prospective sample-size calculation was performed. To assess the amount of information available with the observed sample size, we conducted a simulation in which 92 proteins were tested simultaneously. Power was defined as the probability of detecting at least one truly differential protein while controlling the false discovery rate (FDR) at 5% using the Benjamini-Hochberg method. The correlation structure was estimated from the observed Olink data. Simulations assumed 5, 8, or 10 truly differential proteins, standardized effects (Cohen d) of 0.5, 0.6, 0.7, or 0.8, and total sample sizes ranging from 40 to 200 in increments of 10 (**Fig. 1**). The simulation was intended to characterize the available sample size rather than to define a target sample size.

**Figure 1.**
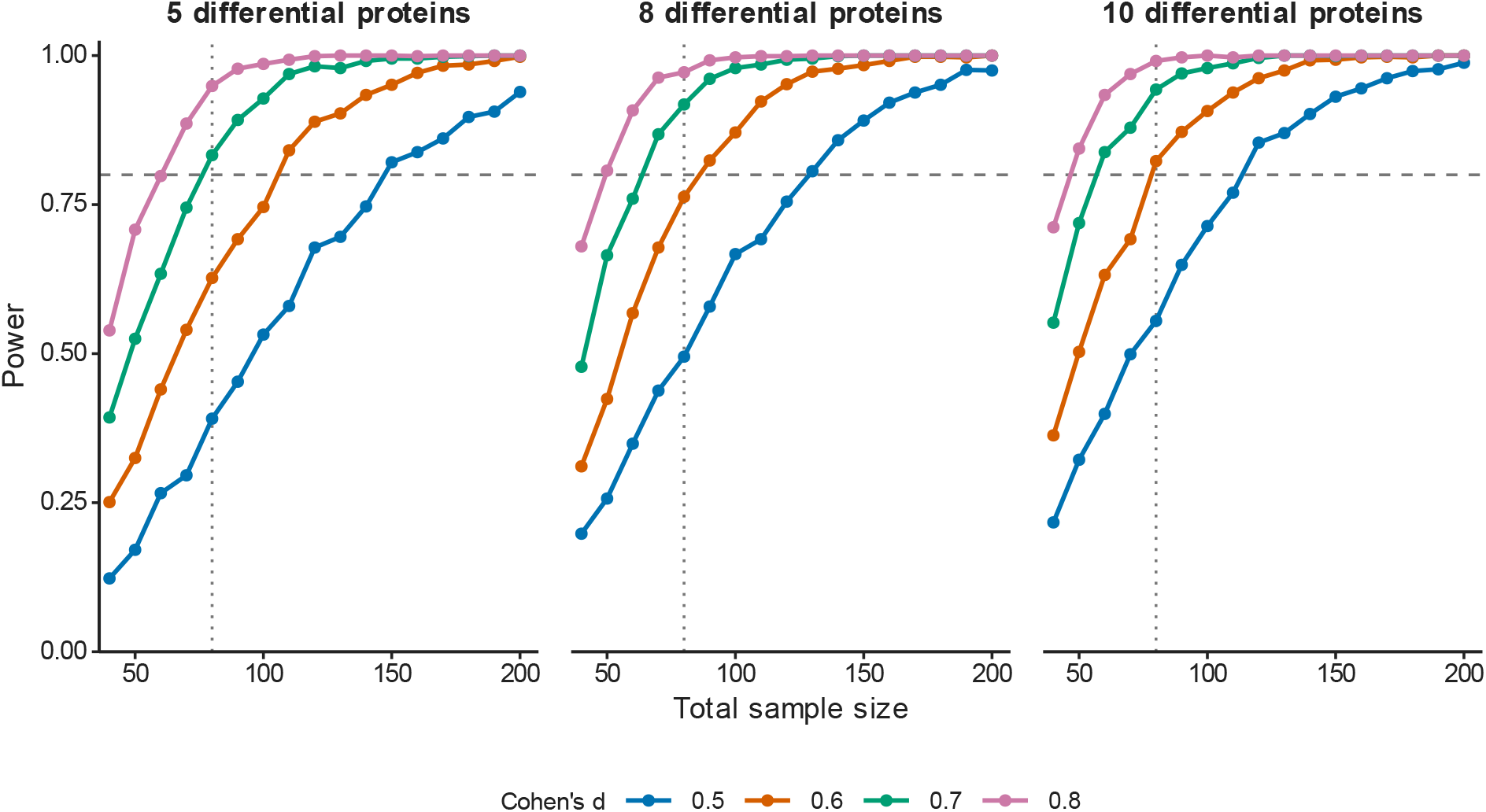
Exploratory simulation of sample size and discovery probability. Power was defined as the probability of detecting at least one truly differential protein after testing 92 proteins and controlling the false discovery rate at 5% with the Benjamini-Hochberg procedure. Panels show assumptions of 5, 8, or 10 truly differential proteins; colors show standardized effects (Cohen d, 0.5 to 0.8). The horizontal dashed line denotes 80% power, and the vertical dotted line denotes the observed sample size of 80. This is the probability of at least one discovery, not the power for an individual protein.

### Statistical analysis

Continuous variables were summarized as mean (standard deviation) or median (interquartile range), as appropriate. Categorical variables were summarized as number (percentage). All tests were two-sided.

Differential protein abundance was assessed using protein-specific linear models and empirical Bayes moderation implemented in limma.[26,27] Empirical Bayes moderation was used to stabilize protein-specific variance estimates by borrowing information across proteins in these small samples. Effect sizes were expressed as log2 fold differences. Benjamini-Hochberg-adjusted P values were calculated within each comparison, with an FDR of less than 0.05 considered statistically significant. Volcano plots display the log2 fold difference on the horizontal axis and the negative log10-transformed unadjusted P value on the vertical axis. For each protein, we also calculated the area under the receiver operating characteristic curve (AUROC).

For descriptive presentation, the first 8 proteins were selected after lexicographic sorting by ascending adjusted P value, ascending unadjusted P value, descending absolute log2 fold difference, and descending univariable AUROC, with each successive criterion used only to break ties. This descriptive ranking was not used to restrict the predictors entered into multivariable models.

To prevent information leakage, the primary multivariable analysis included all 92 proteins in an elasticnet logistic regression without outcome-based feature screening before resampling. Elastic-net regularization was used to accommodate the large number of correlated protein predictors relative to the sample size. Model development and performance estimation used stratified repeated nested cross-validation, with 5 outer folds repeated 5 times and 5 inner folds, separating hyperparameter selection from performance evaluation. Within each inner split, missing values were imputed using medians estimated from the inner training set and applied to the corresponding validation set. The elastic-net mixing and regularization parameters were selected by inner-cross-validation AUROC. For each outer split, imputation parameters were re-estimated from the complete outer training set, the selected model was refitted using the outer training set only, and predictions were generated for the untouched outer test set. Standardization was estimated from training data within each elastic-net fit.

Random forest and gradient boosting were evaluated as sensitivity analyses using the same outer and inner splits. Model-specific hyperparameters were selected within the inner cross-validation. For each model, the 5 outer-test predicted probabilities for each participant were averaged to obtain one out-of-fold prediction. The primary discrimination estimand was the AUROC calculated from these participant-level predictions. Percentile 95% CIs were calculated using 10,000 stratified bootstrap resamples of participants, applied to the fixed participant-level averaged out-of-fold predictions without refitting the models.

For each pairwise comparison, the same leakage-resistant elastic-net pipeline was applied to all 92 proteins. The outer loop used 5 folds repeated 5 times, and the inner loop used up to 4 folds without exceeding the size of the smaller group in the outer training set. Performance was summarized using the participant-averaged out-of-fold AUROC with a stratified bootstrap 95% CI.

To assess potential confounding by specimen matrix, the primary differential abundance analysis and the 3 multivariable models were repeated after excluding participants with secondary hemophagocytic lymphohistiocytosis, the only subgroup assessed using EDTA plasma. The same statistical procedures were used as in the primary analysis. For the all-serum sensitivity analysis, fold- and repeat-level AUROC summaries are presented as secondary diagnostics of resampling stability in Supplementary Table S2. We also repeated the differential abundance analysis and all 3 models after excluding samples flagged as Warning in the laboratory QC report, using the same statistical procedures and refitting the complete nested cross-validation pipeline in the retained participants. Further details of the sensitivity analyses are provided in the Supplementary Methods.

All analyses were performed using R version 4.3.1.

## Results

### Participants

The analysis included 80 participants: 34 with sepsis and 46 with sepsis mimics (Table 1). The sepsis-mimic group comprised 10 participants with COVID-19, 7 with noninfectious ARDS, 8 with aHUS, 11 with secondary HLH, and 10 with TAFRO syndrome. The median (IQR) SOFA score at enrollment was 8.5 (4.2-12) among 34 participants in the sepsis group. The selection of participants from the source cohorts and inclusion in the primary and sensitivity analyses are shown in Supplementary Fig. S2.

**Table 1.**
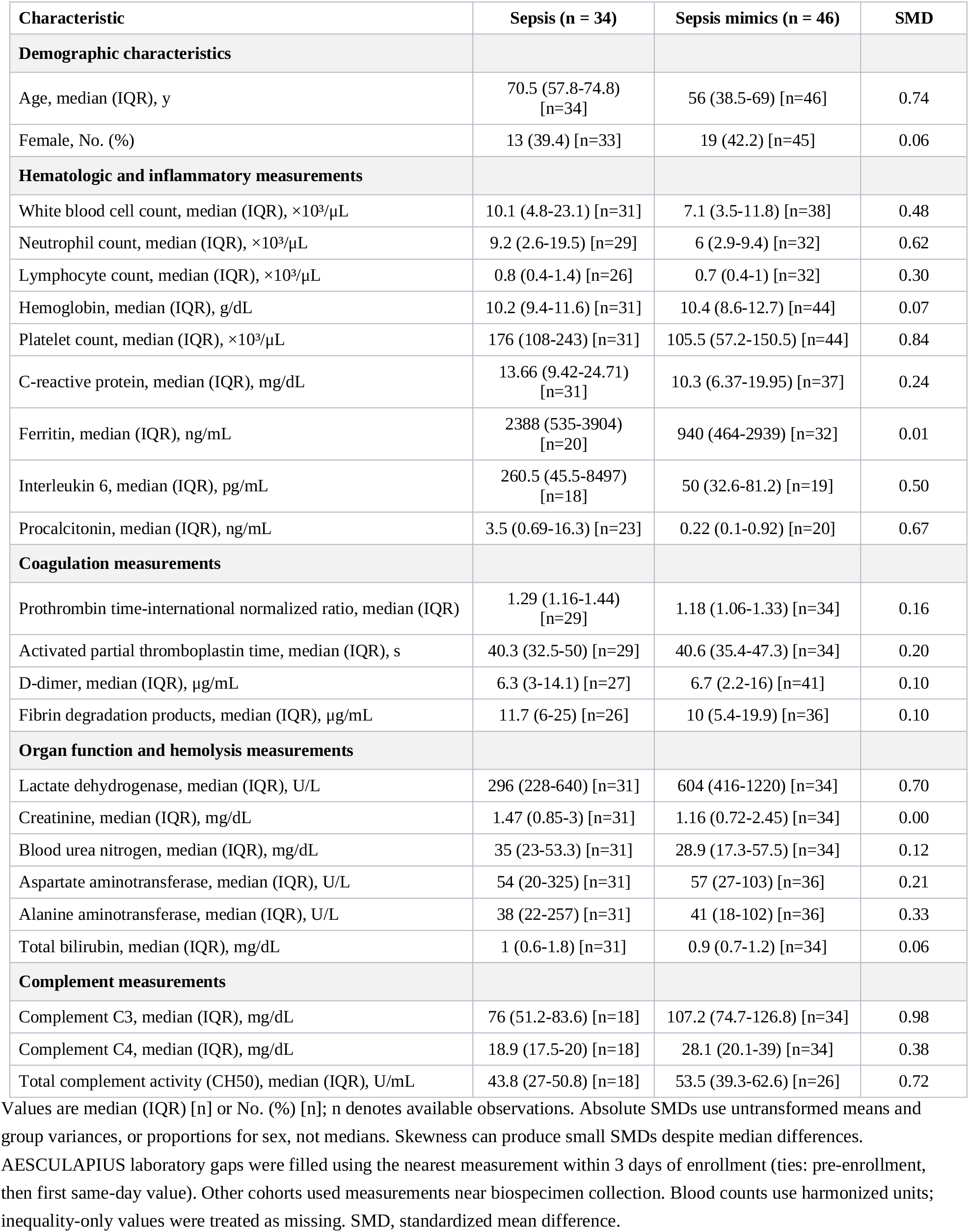
Participant Characteristics and Clinical Laboratory Measurements.

| Characteristic | Sepsis (n = 34) | Sepsis mimics (n = 46) | SMD |
| --- | --- | --- | --- |
| **Demographic characteristics** |  |  |  |
| Age, median (IQR), y | 70.5 (57.8-74.8) [n=34] | 56 (38.5-69) [n=46] | 0.74 |
| Female, No. (%) | 13 (39.4) [n=33] | 19 (42.2) [n=45] | 0.06 |
| **Hematologic and inflammatory measurements** |  |  |  |
| White blood cell count, median (IQR), $\times 10^3/\mu\text{L}$ | 10.1 (4.8-23.1) [n=31] | 7.1 (3.5-11.8) [n=38] | 0.48 |
| Neutrophil count, median (IQR), $\times 10^3/\mu\text{L}$ | 9.2 (2.6-19.5) [n=29] | 6 (2.9-9.4) [n=32] | 0.62 |
| Lymphocyte count, median (IQR), $\times 10^3/\mu\text{L}$ | 0.8 (0.4-1.4) [n=26] | 0.7 (0.4-1) [n=32] | 0.30 |
| Hemoglobin, median (IQR), g/dL | 10.2 (9.4-11.6) [n=31] | 10.4 (8.6-12.7) [n=44] | 0.07 |
| Platelet count, median (IQR), $\times 10^3/\mu\text{L}$ | 176 (108-243) [n=31] | 105.5 (57.2-150.5) [n=44] | 0.84 |
| C-reactive protein, median (IQR), mg/dL | 13.66 (9.42-24.71) [n=31] | 10.3 (6.37-19.95) [n=37] | 0.24 |
| Ferritin, median (IQR), ng/mL | 2388 (535-3904) [n=20] | 940 (464-2939) [n=32] | 0.01 |
| Interleukin 6, median (IQR), pg/mL | 260.5 (45.5-8497) [n=18] | 50 (32.6-81.2) [n=19] | 0.50 |
| Procalcitonin, median (IQR), ng/mL | 3.5 (0.69-16.3) [n=23] | 0.22 (0.1-0.92) [n=20] | 0.67 |
| **Coagulation measurements** |  |  |  |
| Prothrombin time-international normalized ratio, median (IQR) | 1.29 (1.16-1.44) [n=29] | 1.18 (1.06-1.33) [n=34] | 0.16 |
| Activated partial thromboplastin time, median (IQR), s | 40.3 (32.5-50) [n=29] | 40.6 (35.4-47.3) [n=34] | 0.20 |
| D-dimer, median (IQR), $\mu\text{g/mL}$ | 6.3 (3-14.1) [n=27] | 6.7 (2.2-16) [n=41] | 0.10 |
| Fibrin degradation products, median (IQR), $\mu\text{g/mL}$ | 11.7 (6-25) [n=26] | 10 (5.4-19.9) [n=36] | 0.10 |
| **Organ function and hemolysis measurements** |  |  |  |
| Lactate dehydrogenase, median (IQR), U/L | 296 (228-640) [n=31] | 604 (416-1220) [n=34] | 0.70 |
| Creatinine, median (IQR), mg/dL | 1.47 (0.85-3) [n=31] | 1.16 (0.72-2.45) [n=34] | 0.00 |
| Blood urea nitrogen, median (IQR), mg/dL | 35 (23-53.3) [n=31] | 28.9 (17.3-57.5) [n=34] | 0.12 |
| Aspartate aminotransferase, median (IQR), U/L | 54 (20-325) [n=31] | 57 (27-103) [n=36] | 0.21 |
| Alanine aminotransferase, median (IQR), U/L | 38 (22-257) [n=31] | 41 (18-102) [n=36] | 0.33 |
| Total bilirubin, median (IQR), mg/dL | 1 (0.6-1.8) [n=31] | 0.9 (0.7-1.2) [n=34] | 0.06 |
| **Complement measurements** |  |  |  |
| Complement C3, median (IQR), mg/dL | 76 (51.2-83.6) [n=18] | 107.2 (74.7-126.8) [n=34] | 0.98 |
| Complement C4, median (IQR), mg/dL | 18.9 (17.5-20) [n=18] | 28.1 (20.1-39) [n=34] | 0.38 |
| Total complement activity (CH50), median (IQR), U/mL | 43.8 (27-50.8) [n=18] | 53.5 (39.3-62.6) [n=26] | 0.72 |

Values are median (IQR) [n] or No. (%) [n]; n denotes available observations. Absolute SMDs use untransformed means and group variances, or proportions for sex, not medians. Skewness can produce small SMDs despite median differences. AESCULAPIUS laboratory gaps were filled using the nearest measurement within 3 days of enrollment (ties: pre-enrollment, then first same-day value). Other cohorts used measurements near biospecimen collection. Blood counts use harmonized units; inequality-only values were treated as missing. SMD, standardized mean difference.

The simulation indicated that a total sample size of 80 did not reach 80% power when only a few proteins had moderate between-group differences after adjustment for multiple comparisons. Power increased when effect sizes were larger or more proteins were truly differential (**Fig. 1**).

### Primary differential analysis

No protein met the FDR threshold in the comparison between sepsis and the pooled sepsis-mimic group. Tumor necrosis factor receptor superfamily member 9, C-X-C motif chemokine ligand 1, and leukemia inhibitory factor receptor were among the proteins with higher values in the sepsis group (Fig. 2). Mean (SD) NPX for tumor necrosis factor receptor superfamily member 9 was 7.33 (1.33) in sepsis and 6.54 (1.21) in sepsis mimics (unadjusted P = 0.007; FDR-adjusted P = 0.345). The highest single-protein AUROC was 0.676 for C-X-C motif chemokine ligand 1, and the distributions of the 8 leading proteins are shown in Fig. 3. Complete results for all 92 proteins are provided in Supplementary Data 3.

**Figure 2.**
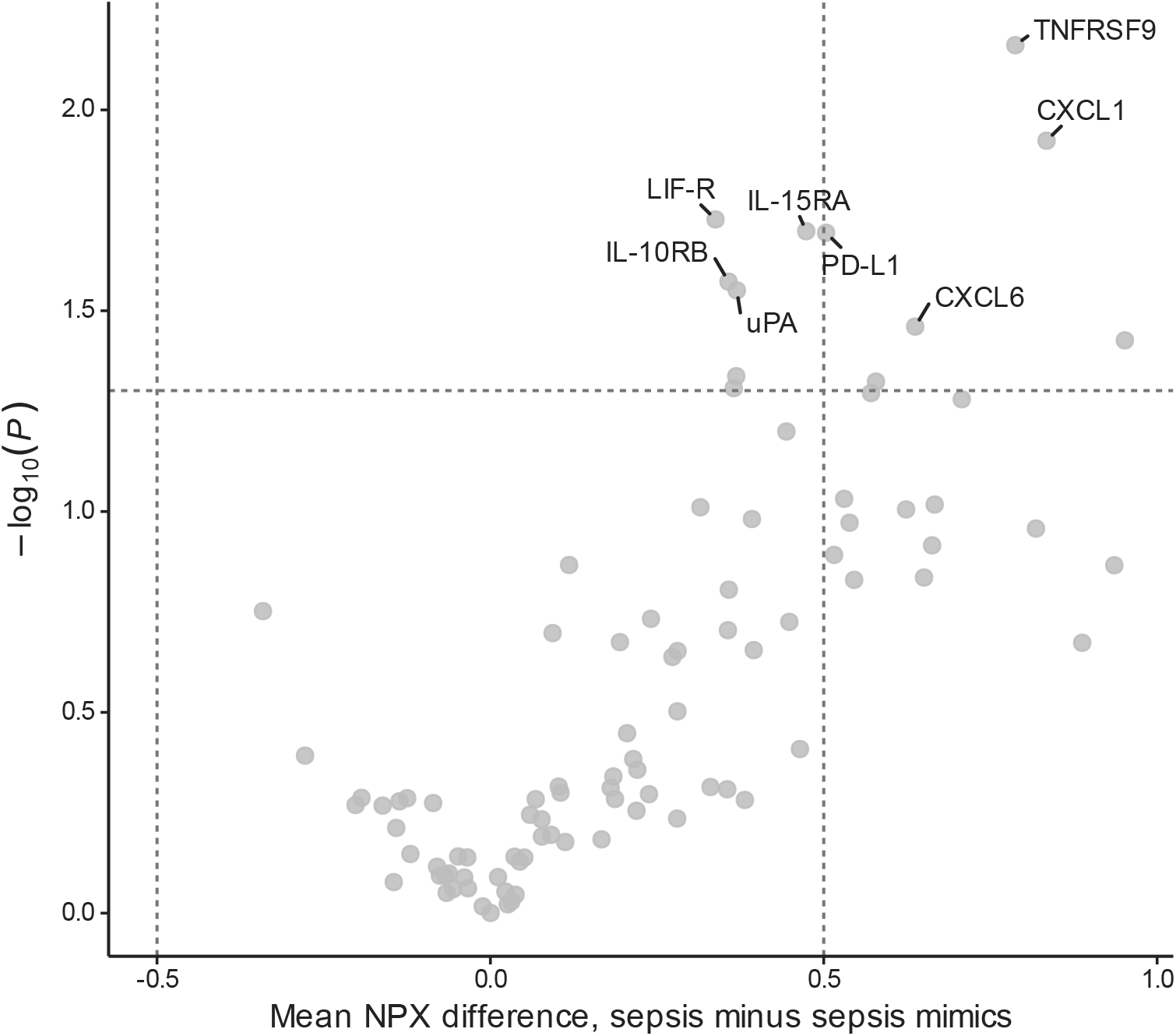
Differential protein abundance between sepsis and the pooled comparator group. The volcano plot shows 92 proteins. The horizontal axis represents the estimated mean Normalized Protein eXpression difference (sepsis minus sepsis mimics) on the log2 Normalized Protein eXpression scale. The vertical axis is the negative log10-transformed unadjusted P value. Vertical dashed lines indicate an absolute difference of 0.5, and the horizontal dashed line indicates an unadjusted P value of 0.05. Labels identify the 8 leading proteins in the descriptive ranking. NPX indicates Normalized Protein eXpression.

**Figure 3.**
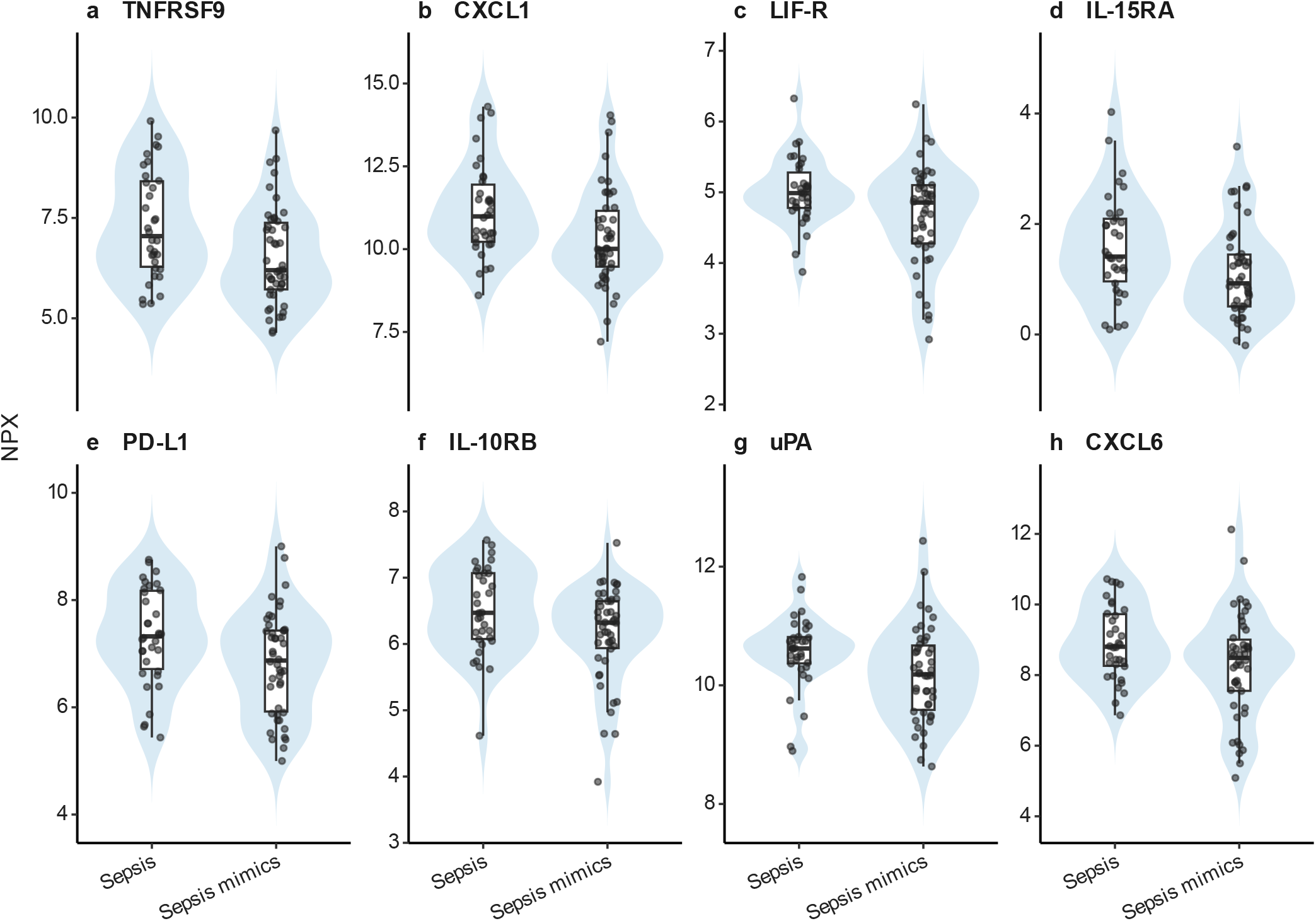
Distributions of the 8 leading proteins. Violin plots show the distributions of Normalized Protein eXpression values, box plots show medians and interquartile ranges, and points show individual participants. CXCL1 indicates C-X-C motif chemokine ligand 1; CXCL6, C-X-C motif chemokine ligand 6; IL-10RB, interleukin-10 receptor subunit beta; IL-15RA, interleukin-15 receptor subunit alpha; LIF-R, leukemia inhibitory factor receptor; NPX, Normalized Protein eXpression; PD-L1, programmed death-ligand 1; TNFRSF9, tumor necrosis factor receptor superfamily member 9; uPA, urokinase-type plasminogen activator. Panels show a, TNFRSF9; b, CXCL1; c, LIF-R; d, IL-15RA; e, PD-L1; f, IL-10RB; g, uPA; and h, CXCL6.

### Exploratory multivariable models

In leakage-resistant analyses using all 92 proteins, the participant-averaged out-of-fold AUROC was 0.442 (95% CI, 0.315-0.574) for elastic-net logistic regression, 0.504 (95% CI, 0.366-0.640) for random forest, and 0.535 (95% CI, 0.410-0.662) for gradient boosting. Discrimination was limited across the 3 model families (Fig. 4).

**Figure 4.**
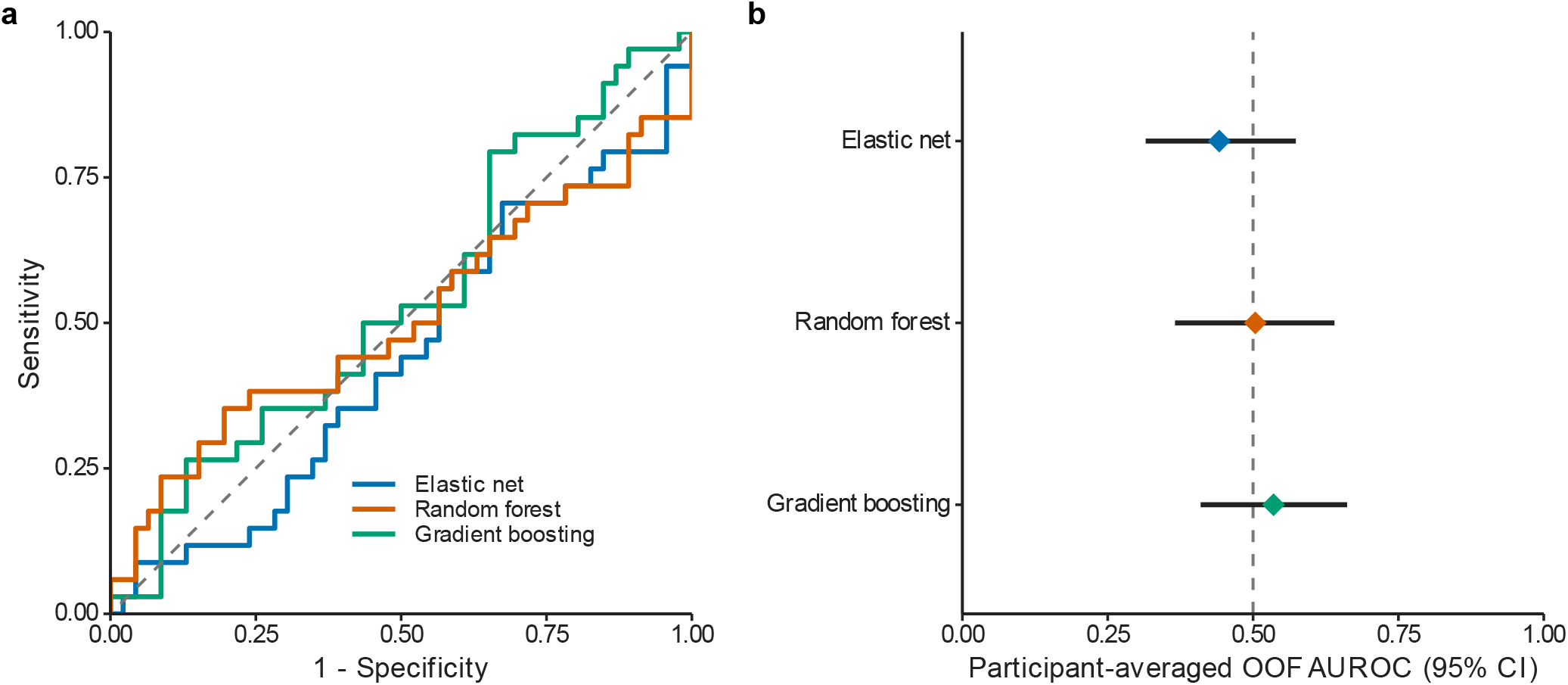
Cross-validated discrimination across 3 all-protein models. a, Empirical receiver operating characteristic curves based on one out-of-fold prediction per participant, calculated by averaging the 5 outer-test probabilities from 5-fold nested cross-validation repeated 5 times. b, Participant-averaged out-of-fold AUROCs with percentile 95% CIs from 10,000 stratified bootstrap resamples of participants. The dashed line denotes an AUROC of 0.50. AUROC indicates area under the receiver operating characteristic curve; CI, confidence interval; and OOF, out of fold.

### Pairwise comparisons

The distribution of protein differences varied among the 5 sepsis-mimic subgroups (**Fig. 5**). More proteins tended to show higher values in sepsis in comparisons with COVID-19, noninfectious ARDS, aHUS, and TAFRO syndrome, whereas more proteins showed lower values in sepsis in the comparison with secondary HLH. None of the pairwise comparisons identified a protein meeting both the FDR threshold and the absolute log2 fold difference threshold of at least 0.5. Complete results for all 92 proteins in each comparison are provided in Supplementary Data 4.

**Figure 5.**
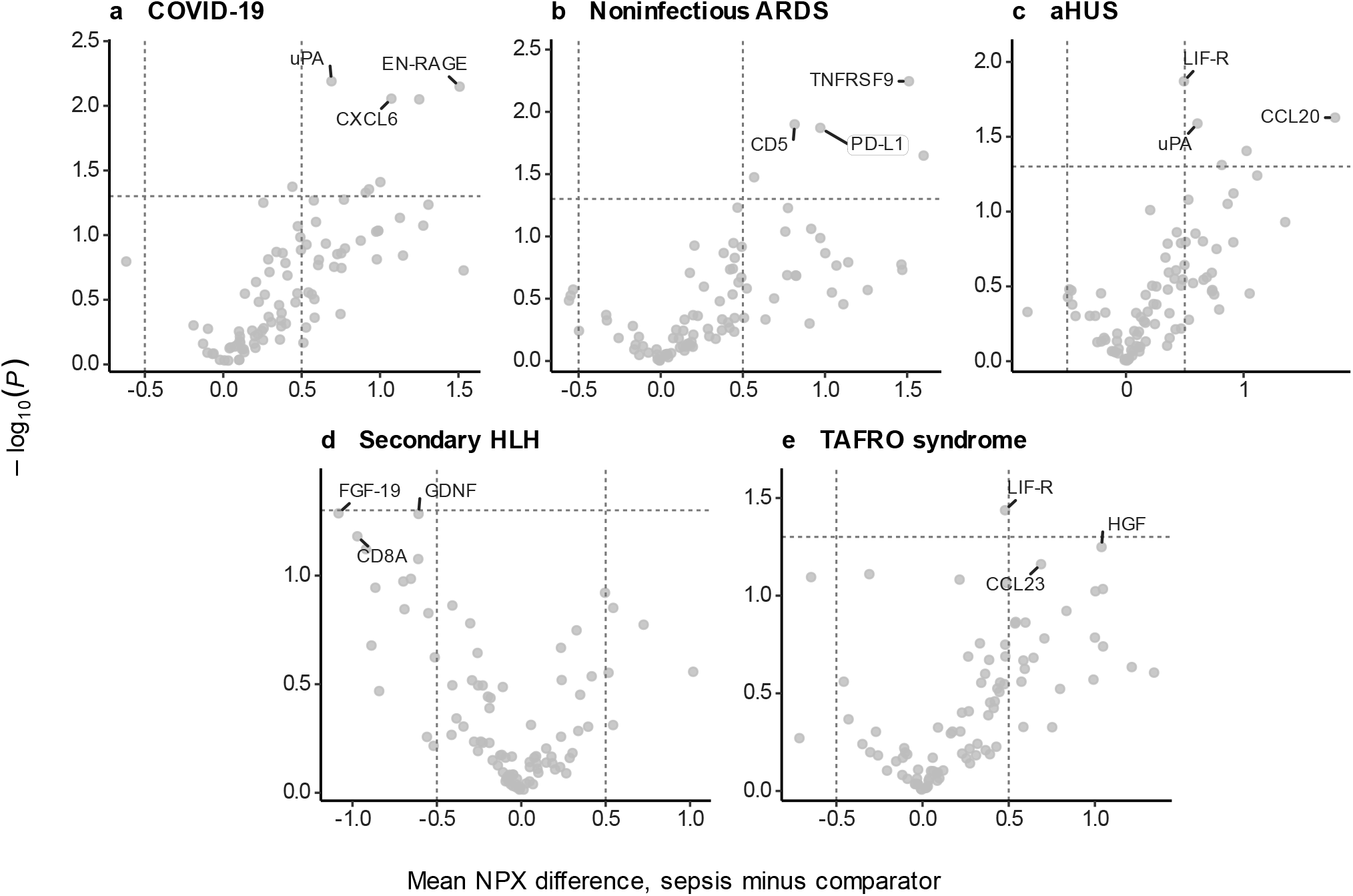
Pairwise differential protein abundance. Volcano plots show comparisons of sepsis with a, coronavirus disease 2019 (n = 10); b, noninfectious acute respiratory distress syndrome (n = 7); c, atypical hemolytic uremic syndrome (n = 8); d, secondary hemophagocytic lymphohistiocytosis (n = 11); and e, thrombocytopenia, anasarca, fever, reticulin fibrosis or renal dysfunction, and organomegaly syndrome (n = 10). The sepsis group included 34 participants in every comparison. The horizontal axis is the estimated mean Normalized Protein eXpression difference (sepsis minus the comparator), and the vertical axis is the negative log10-transformed unadjusted P value. Vertical dashed lines indicate an absolute difference of 0.5, and the horizontal dashed line indicates an unadjusted P value of 0.05.

In pairwise all-protein elastic-net analyses, participant-averaged out-of-fold AUROCs were 0.394 (95% CI, 0.182–0.621) for sepsis vs COVID-19, 0.655 (95% CI, 0.408–0.866) for noninfectious ARDS, 0.415 (95% CI, 0.169–0.669) for aHUS, 0.313 (95% CI, 0.144–0.505) for secondary HLH, and 0.294 (95% CI, 0.112–0.506) for TAFRO syndrome (Fig. 6).

**Figure 6.**
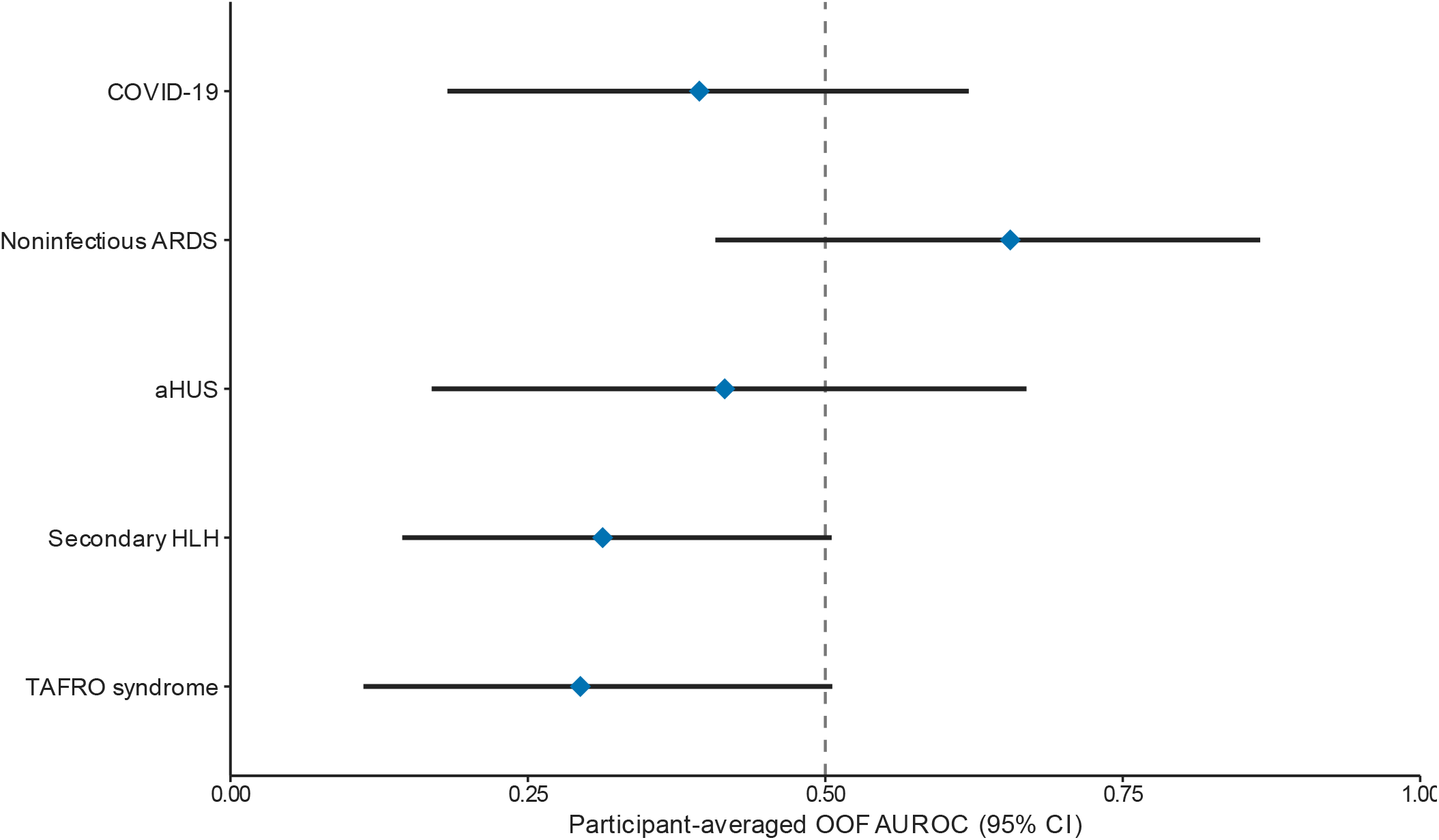
Pairwise elastic-net discrimination across comparator groups. Diamonds indicate participant-averaged out-of-fold AUROCs, and horizontal lines indicate percentile 95% CIs from 10,000 stratified bootstrap resamples of participants. The vertical dashed line denotes an AUROC of 0.50. Each comparison used all 92 proteins in the same elastic-net pipeline. ARDS indicates acute respiratory distress syndrome; aHUS, atypical hemolytic uremic syndrome; AUROC, area under the receiver operating characteristic curve; CI, confidence interval; COVID-19, coronavirus disease 2019; HLH, hemophagocytic lymphohistiocytosis; OOF, out of fold; and TAFRO, thrombocytopenia, anasarca, fever, reticulin fibrosis or renal dysfunction, and organomegaly.

### Sensitivity analyses

In the all-serum sensitivity analysis including 69 participants (34 with sepsis and 35 with other sepsis mimics), participant-averaged out-of-fold AUROC point estimates were higher than those in the primary analysis (0.589–0.603 vs 0.442–0.535), although all 95% CIs included 0.50. No protein met the false discovery rate threshold (Supplementary Tables S1 and S2, Supplementary Fig. S1, and Supplementary Data 1).

Of the 80 study samples, 12 (15%) were flagged as QC Warning. Excluding these samples yielded 68 participants (29 with sepsis and 39 with sepsis mimics) for the QC-Pass sensitivity analysis. AUROCs ranged from 0.489 to 0.534, with all 95% CIs including 0.50; no protein met the FDR threshold (Supplementary Tables S3 and S5, Supplementary Fig. S3, and Supplementary Data 2).

## Discussion

In this study, we investigated whether circulating inflammatory protein profiles could distinguish sepsis from sepsis mimics. No protein met the FDR threshold, and none of the 3 models using all 92 proteins showed clear evidence of discrimination above chance. These findings suggest that a single measurement of an inflammation-focused protein panel alone may be insufficient to distinguish sepsis from sepsis mimics with clinically meaningful and stable accuracy.

One possible explanation is that sepsis and the sepsis mimics included in this study can share systemic inflammation, altered immune responses, and organ dysfunction despite having different initiating causes. [6,28,29] Infection activates innate immune responses to microbial molecules, whereas sterile tissue injury activates overlapping pathways through endogenous molecules released from damaged cells.[28] Immune-mediated inflammatory diseases and toxic reactions can also induce downstream inflammatory responses that overlap with those of sepsis. In addition, the host response in sepsis varies according to the pathogen, site of infection, host characteristics, illness severity, and time from onset. Proinflammatory and immunosuppressive responses do not necessarily occur as separate sequential phases; they may coexist early and change over time.[29,30] Longitudinal immune profiling and plasma proteomic studies have shown that immunological classifications and protein profiles can change during critical illness. [31,32] A single measurement may therefore capture the immune state at the time of sampling without fully distinguishing the evolving pathophysiology of sepsis from that of sepsis mimics.

This interpretation is consistent with the moderate diagnostic performance reported for routine biomarkers. Lengquist and colleagues found that approximately 25% of patients admitted to intensive care with presumed sepsis were subsequently classified as having sepsis mimics. The AUROCs of C-reactive protein and procalcitonin for distinguishing confirmed sepsis from sepsis mimics were 0.71 and 0.70, respectively, whereas the performance of white blood cell count and body temperature was lower. [3] The absence of a clear improvement with multiplex measurement of inflammatory proteins suggests that simply expanding an inflammation panel may not resolve the diagnostic challenge that could not be addressed by conventional inflammatory markers.

At the same time, our results do not argue against host-response-based diagnostic strategies. Palma Medina and colleagues measured 276 proteins related to inflammation, organ injury, immune responses, and coagulation and reported greater discrimination between COVID-19 and sepsis with pneumonia than that achieved with standard clinical variables.[18] In contrast, the present study included a broader range of sepsis mimics and focused on proteins contained in the Olink Target 96 Inflammation panel. The comparatively limited performance observed here may reflect both the biological heterogeneity of the sepsis mimics and the absence of potentially informative signals related to complement activation, coagulation, endothelial injury, metabolism, or tissue injury.

Exploratory pairwise analyses suggested that protein profiles may differ among sepsis-mimic subgroups. However, the higher participant-averaged out-of-fold AUROC for noninfectious ARDS (0.655; 95% CI, 0.408-0.866) was based on only 7 participants and requires independent validation.

The main clinical implication is that an inflammatory protein panel alone is unlikely to serve as a stand-alone test for differentiating sepsis from sepsis mimics. To guide the initiation or continuation of antimicrobial treatment and avoid delays in disease-specific therapies such as immunosuppression or plasma exchange, biomarker results need to be interpreted together with direct evidence of infection, organ-specific findings, hematologic features, imaging, and the clinical course. Future diagnostic strategies should integrate inflammatory proteins with markers of complement activation, coagulation, endothelial injury, and tissue damage, as well as longitudinal changes and clinical information.

This study has several limitations. First, it was an exploratory secondary analysis with a limited overall sample size and very small sepsis-mimic subgroups; the smallest pairwise training partitions contained fewer than 8 comparator observations, limiting the stability of model estimation. Second, sampling times and disease stages were not fully uniform, and a single measurement could not capture temporal changes. Third, the analysis was restricted to proteins included in an inflammation-focused panel and may not have captured other biological pathways relevant to discrimination. Fourth, the use of multiple source cohorts introduced heterogeneity in patient characteristics, sampling, and clinical course. Fifth, EDTA plasma was used exclusively for the HLH cohort, whereas serum was used for all other cohorts; therefore, specimen matrix effects could not be separated from disease- or cohort-related differences in comparisons involving HLH. The higher AUROC point estimates after excluding HLH suggest that discrimination may depend on comparator composition. Sixth, treatment exposure before biospecimen collection was not consistently documented across the parent cohorts. Treatment-related changes in circulating protein levels may have influenced the observed between-group differences and could not be distinguished from disease-related effects. Although all preprocessing and model tuning were confined to training data in the nested-cross-validation analysis, no independent external cohort was available. These limitations should be considered when interpreting the results and planning prospective validation studies.

## Conclusions

Circulating inflammatory protein profiles were insufficient to distinguish sepsis from sepsis mimics with high accuracy in the present cohorts. No single protein met the FDR threshold, and discrimination remained limited across 3 leakage-resistant all-protein models. Distinguishing sepsis from sepsis mimics will likely require diagnostic approaches that integrate disease-specific biological information, longitudinal changes, and clinical data rather than relying on an inflammatory panel alone.

## Data availability

The participant-level data are not publicly available because they contain clinical information from multiple disease-specific and rare-disease cohorts and are governed by the consent and institutional approvals of the parent studies. Requests for access to deidentified data may be directed to the corresponding author and are subject to approval by the relevant institutions and ethics committees. Aggregate protein-level results are provided in Supplementary Data 1–4.

## Code availability

The R scripts used for the statistical analyses and figure generation are available from the corresponding author on request.

## Acknowledgements

This work was supported in part by a Medical Research Grant from the Japan Intractable Diseases Research Foundation (D.K.) and by the Takeda Science Foundation (W.F.). During manuscript preparation, the authors used ChatGPT/Codex (OpenAI) for English translation and language editing. All output was reviewed and revised by the authors, who take full responsibility for the accuracy, integrity, and content of the manuscript.

## Author contributions

D.K. conceived the study, performed the formal analysis, prepared the visualizations, and drafted the manuscript. D.K., A.N., T.Y., N.K., S.M., R.S., S.F., T.K., A.K., W.F., and Y.K. contributed to investigation and resources. H.O. and all other authors contributed to interpretation of the findings and critical revision of the manuscript. All authors reviewed and approved the final version of the manuscript.

## Additional information

Supplementary Information accompanies this article. Correspondence and requests for materials should be addressed to D.K.

## References

1. Singer, M. et al. The Third International Consensus Definitions for Sepsis and Septic Shock (Sepsis-3). JAMA 315, 801–810 (2016). 10.1001/jama.2016.0287

2. Rudd, K. E. et al. Global, regional, and national sepsis incidence and mortality, 1990-2017: analysis for the Global Burden of Disease Study 2017. Lancet 395, 200–211 (2020). 10.1016/S0140-6736(19)32989-7

3. Lengquist, M. et al. Sepsis mimics among presumed sepsis patients at intensive care admission: a retrospective observational study. Infection 52, 1041–1053 (2024). 10.1007/s15010-023-02158-w

4. Litell, J. M., Guirgis, F., Driver, B., Jones, A. E. & Puskarich, M. A. Most emergency department patients meeting sepsis criteria are not diagnosed with sepsis at discharge. Acad. Emerg. Med. 28, 745–752 (2021). 10.1111/acem.14265

5. Long, B. & Koyfman, A. Clinical mimics: an emergency medicine-focused review of sepsis mimics. J. Emerg. Med. 52, 34–42 (2017). 10.1016/j.jemermed.2016.07.102

6. Kellum, J. A., Formeck, C. L., Kernan, K. F., Gomez, H. & Carcillo, J. A. Subtypes and mimics of sepsis. Crit. Care Clin. 38, 195–211 (2022). 10.1016/j.ccc.2021.11.013

7. Story, C. M., Gerber, G. F. & Chaturvedi, S. Medical consult: aHUS, TTP? How to distinguish and what to do. Hematology Am. Soc. Hematol. Educ. Program 2023, 745–753 (2023). 10.1182/hematology.2023000501

8. La Rosee, P. et al. Recommendations for the management of hemophagocytic lymphohistiocytosis in adults. Blood 133, 2465–2477 (2019). 10.1182/blood.2018894618

9. van Rhee, F. et al. International, evidence-based consensus treatment guidelines for idiopathic multicentric Castleman disease. Blood 132, 2115–2124 (2018). 10.1182/blood-2018-07-862334

10. Pierrakos, C., Velissaris, D., Bisdorff, M., Marshall, J. C. & Vincent, J. L. Biomarkers of sepsis: time for a reappraisal. Crit. Care 24, 287 (2020). 10.1186/s13054-020-02993-5

11. Wacker, C., Prkno, A., Brunkhorst, F. M. & Schlattmann, P. Procalcitonin as a diagnostic marker for sepsis: a systematic review and meta-analysis. Lancet Infect. Dis. 13, 426–435 (2013). 10.1016/S1473-3099(12)70323-7

12. Iwase, S. et al. Interleukin-6 as a diagnostic marker for infection in critically ill patients: a systematic review and meta-analysis. Am. J. Emerg. Med. 37, 260–265 (2019). 10.1016/j.ajem.2018.05.040

13. Sweeney, T. E., Shidham, A., Wong, H. R. & Khatri, P. A comprehensive time-course-based multicohort analysis of sepsis and sterile inflammation reveals a robust diagnostic gene set. Sci. Transl. Med. 7, 287ra71 (2015). 10.1126/scitranslmed.aaa5993

14. Assarsson, E. et al. Homogenous 96-plex PEA immunoassay exhibiting high sensitivity, specificity, and excellent scalability. PLoS ONE 9, e95192 (2014). 10.1371/journal.pone.0095192

15. Bossuyt, P. M. et al. STARD 2015: an updated list of essential items for reporting diagnostic accuracy studies. BMJ 351, h5527 (2015). 10.1136/bmj.h5527

16. von Elm, E. et al. The Strengthening the Reporting of Observational Studies in Epidemiology statement: guidelines for reporting observational studies. Lancet 370, 1453–1457 (2007). 10.1016/S0140-6736(07)61602-X

17. Collins, G. S. et al. TRIPOD+AI statement: updated guidance for reporting clinical prediction models that use regression or machine learning methods. BMJ 385, e078378 (2024). 10.1136/bmj-2023-078378

18. Palma Medina, L. M. et al. Targeted plasma proteomics reveals signatures discriminating COVID-19 from sepsis with pneumonia. Respir. Res. 24, 62 (2023). 10.1186/s12931-023-02364-y

19. Yu, J. et al. Severe COVID-19 has a distinct phenotype from bacterial sepsis: a retrospective cohort study in deceased patients. Ann. Transl. Med. 9, 1054 (2021). 10.21037/atm-21-1291

20. Alhazzani, W. et al. Surviving Sepsis Campaign: guidelines on the management of critically ill adults with coronavirus disease 2019 (COVID-19). Crit. Care Med. 48, e440–e469 (2020). 10.1097/CCM.0000000000004363

21. ARDS Definition Task Force et al. Acute respiratory distress syndrome: the Berlin Definition. JAMA 307, 2526–2533 (2012). 10.1001/jama.2012.5669

22. Atypical Hemolytic Uremic Syndrome Clinical Guide Revision Committee. Clinical Guide for Atypical Hemolytic Uremic Syndrome 2023 (in Japanese). (Tokyo Igakusha, 2023). https://jsn.or.jp/academicinfo/report/aHUS_GL2023.pdf

23. Ravelli, A. et al. 2016 classification criteria for macrophage activation syndrome complicating systemic juvenile idiopathic arthritis: a European League Against Rheumatism/American College of Rheumatology/Paediatric Rheumatology International Trials Organisation collaborative initiative. Arthritis Rheumatol. 68, 566–576 (2016). 10.1002/art.39332

24. Masaki, Y. et al. Proposed diagnostic criteria, disease severity classification and treatment strategy for TAFRO syndrome, 2015 version. Int. J. Hematol. 103, 686–692 (2016). 10.1007/s12185-016-1979-1

25. Masaki, Y. et al. 2019 updated diagnostic criteria and disease severity classification for TAFRO syndrome. Int. J. Hematol. 111, 155–158 (2020). 10.1007/s12185-019-02780-1

26. Smyth, G. K. Linear models and empirical Bayes methods for assessing differential expression in microarray experiments. Stat. Appl. Genet. Mol. Biol. 3, Article 3 (2004). 10.2202/1544-6115.1027

27. Ritchie, M. E. et al. limma powers differential expression analyses for RNA-sequencing and microarray studies. Nucleic Acids Res. 43, e47 (2015). 10.1093/nar/gkv007

28. Chen, G. Y. & Nunez, G. Sterile inflammation: sensing and reacting to damage. Nat. Rev. Immunol. 10, 826–837 (2010). 10.1038/nri2873

29. van der Poll, T., Shankar-Hari, M. & Wiersinga, W. J. The immunology of sepsis. Immunity 54, 2450–2464 (2021). 10.1016/j.immuni.2021.10.012

30. Rubio, I. et al. Current gaps in sepsis immunology: new opportunities for translational research. Lancet Infect. Dis. 19, e422–e436 (2019). 10.1016/S1473-3099(19)30567-5

31. Balch, J. A. et al. Defining critical illness using immunological endotypes in patients with and without sepsis: a cohort study. Crit. Care 27, 292 (2023). 10.1186/s13054-023-04571-x

32. Mi, Y. et al. High-throughput mass spectrometry maps the sepsis plasma proteome and differences in patient response. Sci. Transl. Med. 16, eadh0185 (2024). 10.1126/scitranslmed.adh0185

