## Supplementary Information for "Inflammatory protein profiling shows limited ability to distinguish sepsis from sepsis mimics"

### Supplementary Methods

##### All-serum sensitivity analysis

To assess potential confounding by specimen matrix, we repeated the primary analyses after excluding the 11 participants with secondary hemophagocytic lymphohistiocytosis, the only subgroup assessed using EDTA plasma. The all-serum analysis therefore included 69 participants: 34 with sepsis and 35 with other sepsis mimics (10 with coronavirus disease 2019, 7 with noninfectious acute respiratory distress syndrome, 8 with atypical hemolytic uremic syndrome, and 10 with thrombocytopenia, anasarca, fever, reticulin fibrosis or renal dysfunction, and organomegaly syndrome).

Differential protein abundance was assessed for all 92 proteins using protein-specific linear models with empirical Bayes moderation, and P values were adjusted using the Benjamini-Hochberg procedure. The elastic-net logistic regression, random forest, and gradient boosting models were evaluated using the same nested cross-validation framework as in the primary analysis: a 5-fold outer loop repeated 5 times and 5-fold inner cross-validation for hyperparameter selection. All 92 proteins were retained, and median imputation was estimated separately within each training set. The 5 repeated outer-test probabilities were averaged for each participant, and the primary discrimination estimand was the AUROC calculated from these participant-level predictions. Percentile 95% CIs were calculated using 10,000 stratified bootstrap resamples of participants, applied to the fixed participant-level averaged out-of-fold predictions without refitting the models.

##### QC-Pass sensitivity analysis

Of the 80 study samples, 12 (15%) were flagged as QC Warning in the laboratory report: 5 from participants with sepsis, 2 with COVID-19, 2 with noninfectious ARDS, 1 with secondary HLH, and 2 with TAFRO syndrome. All 80 samples were retained in the primary analysis. Exclusion of the 12 flagged samples yielded 68 participants, comprising 29 with sepsis and 39 with sepsis mimics (8 with COVID-19, 5 with noninfectious ARDS, 8 with aHUS, 10 with secondary HLH, and 8 with TAFRO syndrome). This subset was defined by sample-level QC status, independently of the all-serum restriction.

All 92 proteins were reassessed using the same linear models with empirical Bayes moderation and Benjamini–Hochberg adjustment. The 3 models were refitted using 5-fold outer cross-validation repeated 5 times and 5-fold inner cross-validation, with all preprocessing and hyperparameter selection confined to the respective training sets. AUROCs were calculated from participant-averaged outer-test predictions. Percentile 95% CIs used 10,000 stratified participant-level bootstrap resamples of these fixed predictions, without refitting models within the bootstrap.

##### Supplementary Table S1. Leading differential protein results in the all-serum sensitivity analysis

Proteins are ranked by false discovery rate-adjusted P value and then unadjusted P value. The complete results for all 92 proteins are provided in Supplementary Data 1.

| Protein | Mean NPX, sepsis | Mean NPX, mimics | Mean difference | P value | FDR-adjusted P value | AUROC |
|---|---|---|---|---|---|---|
| uPA | 10.56 | 10.01 | 0.545 | 0.002 | 0.091 | 0.729 |
| TNFRSF9 | 7.33 | 6.35 | 0.979 | 0.002 | 0.091 | 0.717 |
| LIF-R | 5.01 | 4.58 | 0.428 | 0.007 | 0.128 | 0.683 |
| CCL20 | 9.22 | 7.94 | 1.280 | 0.009 | 0.128 | 0.676 |
| CD5 | 5.88 | 5.36 | 0.516 | 0.009 | 0.128 | 0.685 |
| CXCL1 | 11.17 | 10.23 | 0.934 | 0.009 | 0.128 | 0.703 |
| PD-L1 | 7.32 | 6.71 | 0.615 | 0.010 | 0.128 | 0.673 |
| CCL19 | 12.34 | 11.40 | 0.945 | 0.015 | 0.171 | 0.685 |
| EN-RAGE | 6.79 | 5.86 | 0.927 | 0.022 | 0.184 | 0.667 |
| CSF-1 | 10.31 | 10.12 | 0.190 | 0.023 | 0.184 | 0.652 |

AUROC indicates area under the receiver operating characteristic curve; FDR, false discovery rate; and NPX, Normalized Protein eXpression. Mean difference is sepsis minus sepsis mimics on the log2 NPX scale.

##### Supplementary Table S2. Multivariable model performance in the all-serum sensitivity analysis

The participant-averaged out-of-fold AUROC was the primary discrimination estimand. Mean outer-fold, median outer-fold, and mean repeat-level AUROCs are shown only as secondary diagnostics of resampling stability.

| Model | Participant-averaged OOF AUROC (95% CI) | Mean outer-fold AUROC (SD) | Median outer-fold AUROC (range) | Mean repeat-level AUROC (SD) |
|---|---|---|---|---|
| Elastic-net logistic regression | 0.603 (0.465-0.737) | 0.602 (0.137) | 0.592 (0.429-0.857) | 0.611 (0.047) |
| Random forest | 0.597 (0.459-0.729) | 0.594 (0.091) | 0.592 (0.429-0.755) | 0.592 (0.031) |
| Gradient boosting | 0.589 (0.450-0.724) | 0.593 (0.096) | 0.633 (0.408-0.738) | 0.587 (0.025) |

AUROC indicates area under the receiver operating characteristic curve; CI, confidence interval; OOF, out of fold; and SD, standard deviation. The 95% CI was obtained from 10,000 stratified participant-level bootstrap resamples.

**Supplementary Table S3. Multivariable model performance in the QC-Pass sensitivity analysis**

| Model | AUROC | 95% CI |
|---|---|---|
| Elastic-net logistic regression | 0.523 | 0.379–0.661 |
| Random forest | 0.534 | 0.385–0.683 |
| Gradient boosting | 0.489 | 0.332–0.644 |

The analysis included 29 participants with sepsis and 39 with sepsis mimics. AUROC indicates area under the receiver operating characteristic curve; CI, confidence interval; and QC, quality control.

**Supplementary Table S4. Laboratory assay performance for the 88-sample analytical batch**

| Metric | Value |
|---|---|
| Samples in the analytical batch, No. | 88 |
| Samples included in the present study, No. | 80 |
| Analytical plates, No. | 1 |
| Batch samples passing sample-level QC, No./total (%) | 76/88 (86) |
| Study samples passing sample-level QC, No./total (%) | 68/80 (85) |
| Proteins detected in >75% of batch samples, No./total (%) | 75/92 (82) |
| Mean intra-assay CV, % | 3 |
| Assays with available intra-assay CV estimates, No./total | 68/92 |
| Assays with intra-assay CV ≥15%, No./evaluable assays | 0/68 |
| Assays with unavailable intra-assay CV estimates, No./total | 24/92 |
| Inter-assay CV | Not assessed |

Metrics were taken from the original laboratory report for the full analytical batch, which included the 80 study samples. Detection frequency was reported for the batch rather than recalculated for the study subset. Intra-assay CVs were calculated using pooled plasma controls on the linear NPX scale. All 68 assays with available CV estimates were below 15%; estimates were unavailable for 24 assays. Inter-assay variability was not assessed because all samples were analyzed on a single plate. CV indicates coefficient of variation; NPX, Normalized Protein eXpression; and QC, quality control.

**Supplementary Table S5. Leading differential protein results in the QC-Pass sensitivity analysis**

Proteins are ranked by ascending FDR-adjusted P value and then ascending unadjusted P value. The analysis included 29 participants with sepsis and 39 with sepsis mimics. Complete results for all 92 proteins are provided in Supplementary Data 2.

| Protein | Mean NPX, sepsis | Mean NPX, mimics | Mean difference | P value | FDR-adjusted P value | AUROC |
|---|---|---|---|---|---|---|
| TNFRSF9 | 7.51 | 6.57 | 0.945 | 0.003 | 0.259 | 0.694 |
| IL-10RB | 6.60 | 6.11 | 0.494 | 0.006 | 0.259 | 0.698 |
| LIF-R | 5.03 | 4.61 | 0.415 | 0.009 | 0.259 | 0.663 |
| CXCL6 | 8.88 | 8.09 | 0.786 | 0.020 | 0.259 | 0.664 |
| LAP TGF-beta-1 | 7.01 | 6.54 | 0.467 | 0.021 | 0.259 | 0.658 |
| CD5 | 5.93 | 5.46 | 0.472 | 0.022 | 0.259 | 0.654 |
| VEGFA | 12.39 | 11.65 | 0.741 | 0.027 | 0.259 | 0.652 |
| uPA | 10.57 | 10.18 | 0.397 | 0.028 | 0.259 | 0.664 |
| IL-15RA | 1.62 | 1.14 | 0.482 | 0.030 | 0.259 | 0.650 |
| IL-1 alpha | 0.81 | 0.09 | 0.721 | 0.032 | 0.259 | 0.615 |

Mean difference is sepsis minus sepsis mimics on the log2 NPX scale. AUROC is descriptive univariable discrimination, not cross-validated multivariable performance. AUROC indicates area under the receiver operating characteristic curve; FDR, false discovery rate; NPX, Normalized Protein eXpression; and QC, quality control.

**Supplementary Figure S1. Discrimination across 3 all-protein models in the all-serum sensitivity analysis**

a, Empirical receiver operating characteristic curves based on one out-of-fold prediction per participant, calculated by averaging the 5 outer-test probabilities from 5-fold nested cross-validation repeated 5 times. b, Participant-averaged out-of-fold AUROCs with percentile 95% CIs from 10,000 stratified bootstrap resamples of participants. AUROCs were 0.603 (95% CI, 0.465-0.737) for elastic net, 0.597 (95% CI, 0.459-0.729) for random forest, and 0.589 (95% CI, 0.450-0.724) for gradient boosting. The vertical dashed line denotes an AUROC of 0.50. The analysis included 69 participants (34 with sepsis and 35 with other sepsis mimics).

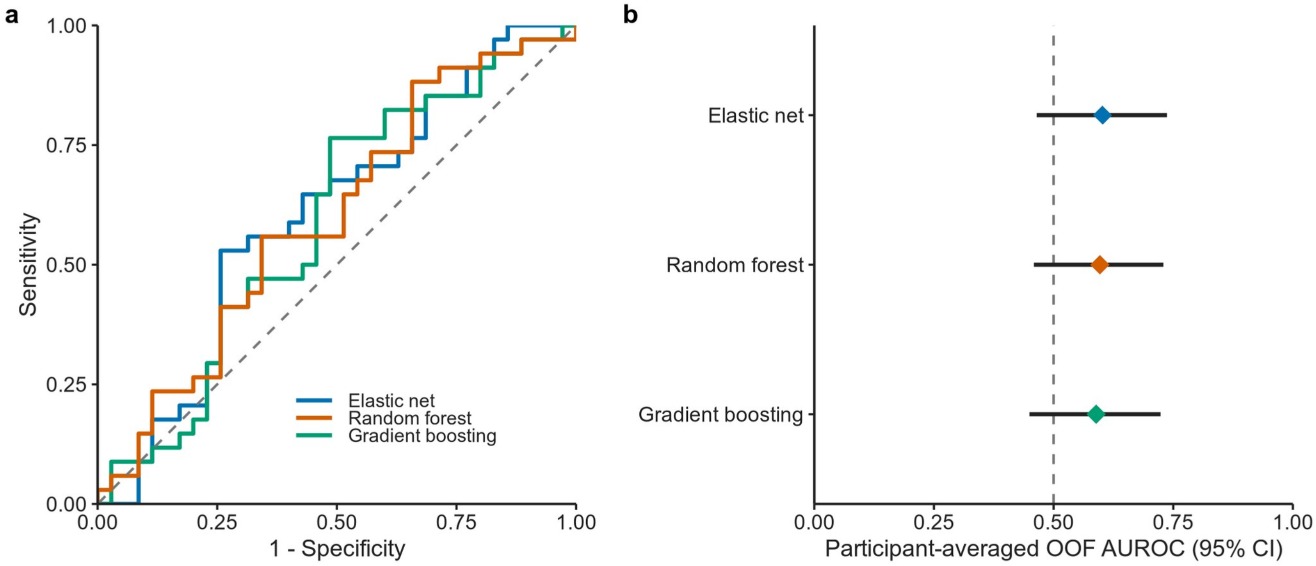

**Supplementary Figure S2. Participant selection and analysis flow**

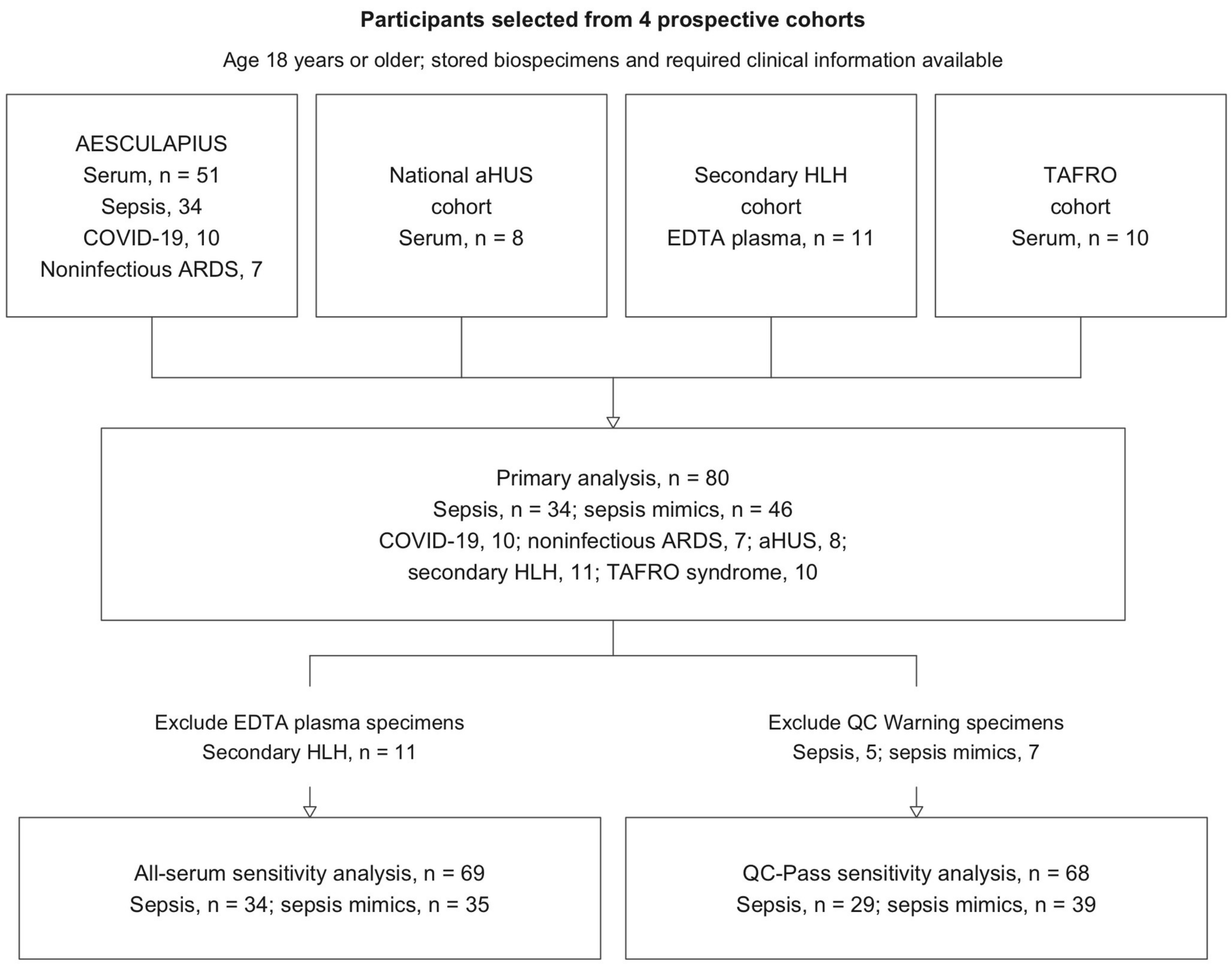


Participants were selected according to the availability of stored biospecimens and the clinical information required for this secondary analysis. The diagram starts with the selected participants; parent-cohort screening denominators and reasons for nonselection are not shown. The primary analysis included 80 participants. Exclusion of the 11 participants with secondary HLH, whose specimens were EDTA plasma, yielded 69 participants for the all-serum sensitivity analysis. Separately, exclusion of 12 samples with QC warnings yielded 68 participants for the QC-Pass sensitivity analysis. QC indicates quality control; ARDS, acute respiratory distress syndrome; aHUS, atypical hemolytic uremic syndrome; COVID-19, coronavirus disease 2019; HLH, hemophagocytic lymphohistiocytosis; and TAFRO, thrombocytopenia, anasarca, fever, reticulin fibrosis or renal dysfunction, and organomegaly.

**Supplementary Figure S3. Discrimination across 3 all-protein models in the QC-Pass sensitivity analysis**

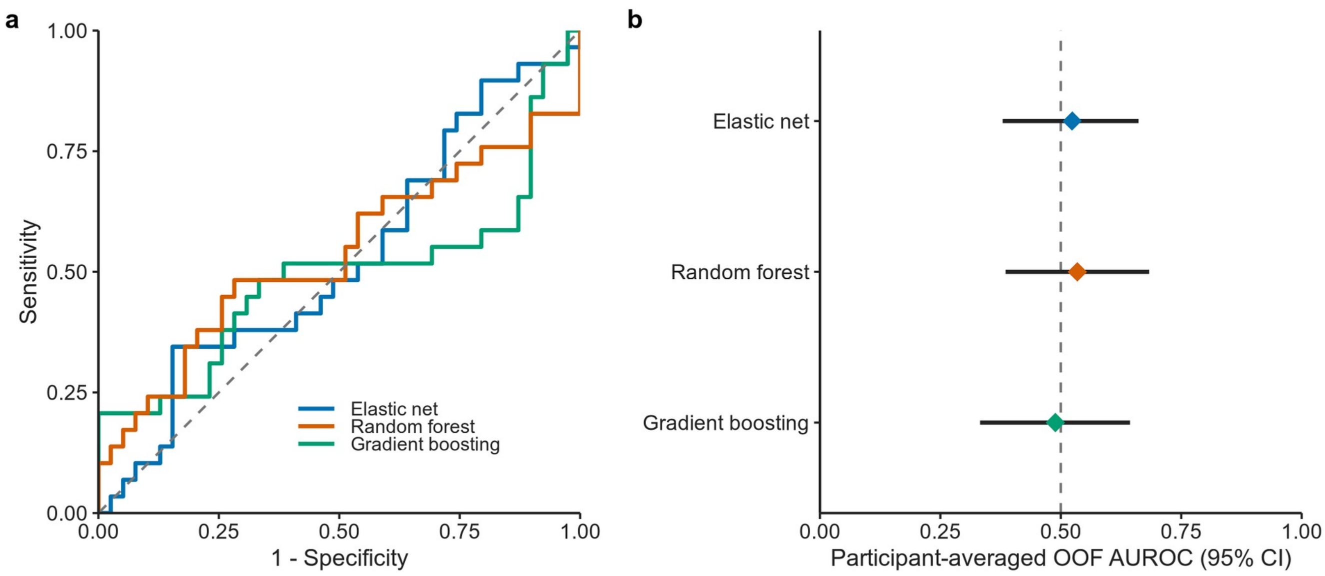


a, Empirical receiver operating characteristic curves based on participant-averaged outer-test predictions. b, Participant-averaged out-of-fold AUROCs with percentile 95% CIs from 10,000 stratified participant-level bootstrap resamples. The analysis included 68 participants (29 with sepsis and 39 with sepsis mimics) after excluding samples with QC warnings. The dashed lines indicate chance discrimination. AUROC indicates area under the receiver operating characteristic curve; CI, confidence interval; OOF, out of fold; and QC, quality control.

#### Supplementary Data

Supplementary Data 1 contains the complete protein-level results for all 92 proteins in the all-serum sensitivity analysis, including group-specific sample sizes and mean Normalized Protein eXpression values, mean differences, unadjusted and false discovery rate-adjusted P values, and univariable areas under the receiver operating characteristic curve.

Supplementary Data 2 contains the complete results for all 92 proteins in the QC-Pass sensitivity analysis, including group-specific sample sizes, mean (SD) NPX values, mean differences, unadjusted and FDR-adjusted P values, and univariable AUROCs.

Supplementary Data 3 contains the complete primary-analysis results for all 92 proteins in the 80 participants (34 with sepsis and 46 with sepsis mimics), including mean NPX values, differences, unadjusted and FDR-adjusted P values, and descriptive univariable AUROCs.

Supplementary Data 4 contains the complete results for all 92 proteins in each of the 5 pairwise comparisons (460 protein–comparison results). Comparators are ordered as COVID-19, noninfectious ARDS, aHUS, secondary HLH, and TAFRO syndrome. FDR adjustment was applied separately within each comparison.
